# How the COVID-19 pandemic is shaping research in Africa: inequalities in scholarly output and collaborations and new opportunities for scientific leadership

**DOI:** 10.1101/2021.04.24.21256053

**Authors:** Maxime Descartes Mbogning Fonkou, Nicola Luigi Bragazzi, Emmanuel Kagning Tsinda, Yagai Bouba, Gideon Sadikiel Mmbando, Jude Dzevela Kong

## Abstract

**Background:** Scientometrics enables scholars to assess and visualize emerging research trends and hot-spots in the scientific literature from a quantitative standpoint. In the last decades, Africa has nearly doubled its absolute count of scholarly output, even though its share in global knowledge production has dramatically decreased. This limited contribution of African scholars to the global research output is in part impacted by the availability of adequate infrastructures and research collaborative networks. The still ongoing COVID-19 pandemic has profoundly impacted the way scholarly research is conducted, published and disseminated. However, the COVID-19 related research focus, the scientific productivity and the research collaborative network of African researchers during the ongoing COVID-19 pandemic remain to be elucidated yet.

**Methods:** This study aimed to clarify the COVID-19 research patterns among African researchers and estimate the strength of collaborations and partnerships between African researchers and scholars from the rest of the world during the COVID-19 pandemic, collecting data from electronic scholarly databases such as Web of Sciences (WoS), PubMed/MEDLINE and African Journals OnLine (AJOL), the largest and prominent platform of African-published scholarly journals.

**Results:** In the present bibliometric study, we found that COVID-19 related collaboration patterns varied among African regions, being shaped and driven by historical, social, cultural, linguistic, and even religious determinants. For instance, most of the scholarly partnerships occurred with formerly colonial countries (like European or North-American countries). In other cases, scholarly ties of North African countries were above all with the Kingdom of Saudi Arabia. In terms of amount of publications, South Africa and Egypt were among the most productive countries.

**Discussion:** Bibliometrics and, in particular, scientometrics can help scholars identify research areas of particular interest, as well as emerging topics, such as the COVID-19 pandemic. With a specific focus on the still ongoing viral outbreak, they can assist decision- and policy-makers in allocating funding and economic-financial, logistic, organizational, and human resources, based on the specific gaps and needs of a given country or research area.

**Conclusions:** In conclusion, the ongoing COVID-19 pandemic has exerting a subtle, complex impact on research and publishing patterns in African countries. On the one hand, it has distorted and even amplified existing inequalities and disparities in terms of amount of scholarly output, share of global knowledge, and patterns of collaborations. On the other hand, COVID-19 provided new opportunities for research collaborations.

## Background

Scientometrics is emerging as a highly specialized branch of information science and as a sub-field of bibliometrics. It enables scholars to assess and visualize emerging research trends and hot-spots in the scientific literature from a quantitative standpoint. Moreover, scientometrics allows a rigorous analysis of patterns in terms of article usage and citations, generating an extensive series of measurements and indicators that can provide policy- and decision-makers with useful information concerning the effectiveness of their policies (1)(2)(3).

In the last decades, Africa has nearly doubled its absolute count of scholarly output (4), even though its share in global knowledge production has dramatically decreased (5), with African countries losing approximately 11% of their share since their peak in 1987, and with Sub-Saharan Africa severely lagging behind and reporting a loss up to 31%. According to some updated statistics (6), African countries contribute to less than 1-1.5% of the global research output (7). This limited contribution of African scholars to the global research output is in part impacted by the availability of adequate infrastructures and research collaborative networks.

The still ongoing “Coronavirus Disease 2019” (COVID-19) pandemic, caused by the emerging “Severe Acute Respiratory Syndrome-related Coronavirus type 2” (SARS-CoV-2), is an unprecedented infectious outbreak. Besides imposing a dramatic toll of cases and deaths, and being devastating both from a societal and economic-financial perspective, COVID-19 has profoundly impacted the way scholarly research is conducted, published and disseminated. Some authors (8) retrieved a pool of 441 articles relevant to the COVID-19 pandemic, approximately half of which (44.90%) were produced by mainland China, followed by the USA, Italy, Germany, and South Korea. Lower-middle-income and low-income countries contributed to 2.95%, and 0.23% of the output, respectively, with a negligible contribution from African countries and territories.

Bibliometric and scientometric analyses have been conducted to explore the emerging research focus related to COVID-19. Such research focus identified by researchers in mid-high income countries include available treatment options, like approved drugs or vaccines, or candidate management strategies (9; 10; 11). While some bibliometric papers focus on summarizing research foci, other scientometric publication have assessed the scholarly output of researchers mainly from countries in Asia, America or Europe (12)(13)(14). However, the COVID-19 related research focus, the scientific productivity and the research collaborative network of African researchers during the ongoing COVID-19 pandemic remain to be elucidated yet.

Therefore, this study aimed to clarify the COVID-19 research patterns among African researchers and estimate the strength of collaborations and partnerships between African researchers and scholars from the rest of the world during the COVID-19 pandemic.

## Methods

The document selection process was as follows.

### Bibliographic search and document identification

To identify the scientific literature on COVID-19 produced in Africa, we used a search string which consisted of terms related to COVID-19, the names of African countries and the main cities of these countries and territories (supplementary data 1) (available here via this link https://github.com/descartesmbogning/How-the-COVID-19-pandemic-is-shaping-research-in-Africa-inequalities-in-scholarly-output-and-collab.git. Data was collected from electronic scholarly databases such as Web of Sciences (WoS), PubMed/MEDLINE and African Journals OnLine (AJOL), the largest and prominent platform of African-published scholarly journals. A database search was made on 12 March 2021 and publication date of papers was restricted to the period between 2019 and 2021. The number of records identified from Pubmed/MEDLINE, WoS and AJOL were 4256, 5,591 and 197, respectively. Figure 1 presents a flow-chart showing the selection process for the documents retained and analyzed in the present study.

**Figure 1:**
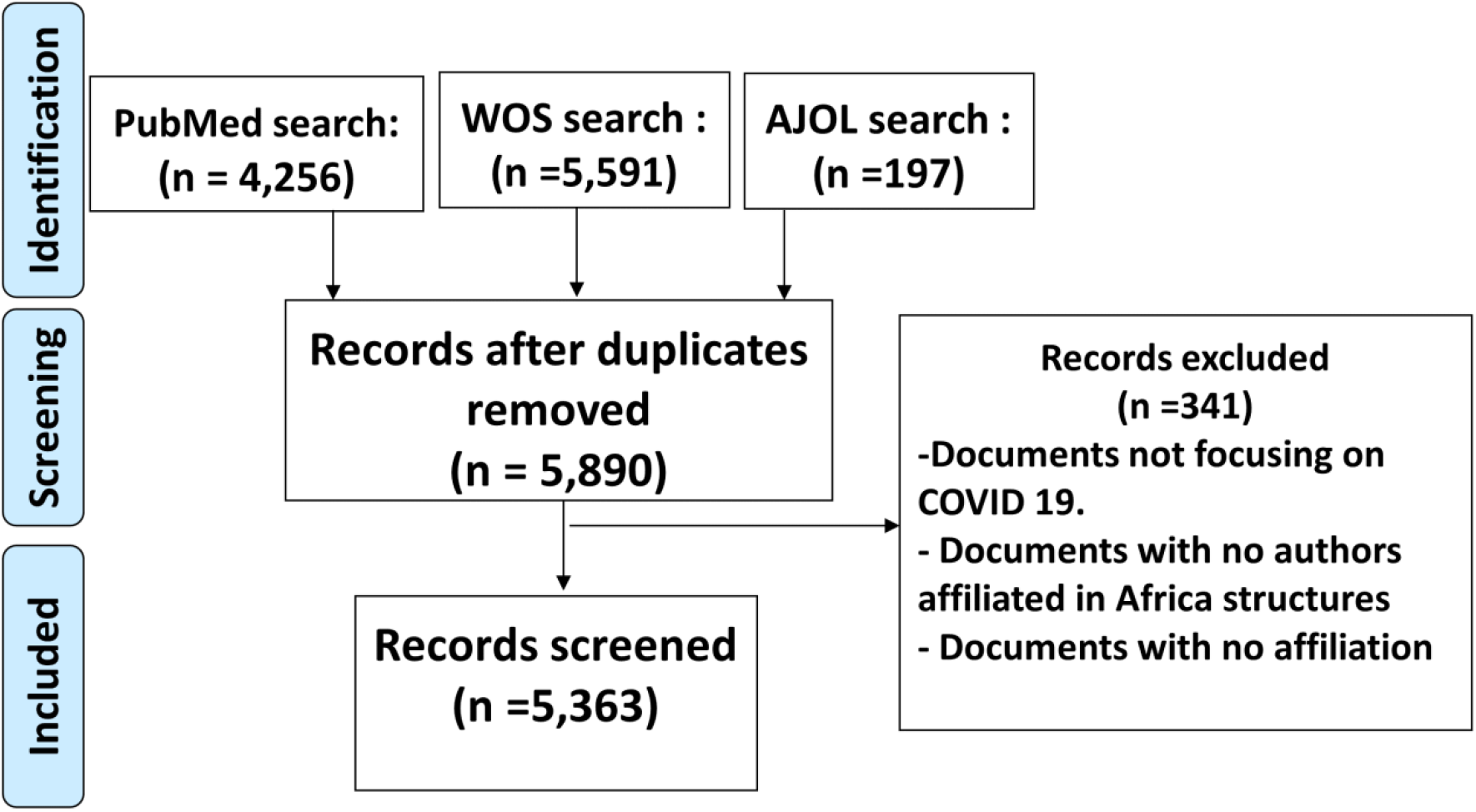
Flow-chart showing the selection process of documents included in the present study

### Download of bibliographic information and review of the quality and standardization of data

Following the bibliographic search and document identification, we downloaded the data from the databases. 5,890 duplicate records were removed and 5,363 records were included for downstream analyses. Duplicates removal was done using ad hoc software (Endnote). The data file was then exported into a Microsoft Office Excel spreadsheet to count and exclude duplicated entries in some bibliographic fields. We found duplicated elements in institutional affiliations. We also reviewed and standardized entries of some fields. For example, among records from WoS, entries with a geographical origin that included “England”, “Scotland”, “Wales”, and “North Ireland” were renamed to “United Kingdom”.

## Data Analyses

To analyze the COVID-19 publications from Africa, we grouped all countries according to World Bank geographical regions (15) and we assigned each country to its corresponding World Bank region. These World Bank regions included: East Asia and Pacific, Europe and Central Asia, Latin America & the Caribbean, Middle East and North Africa (Middle East/North Africa), North America, South Asia, Sub-Saharan Africa (Eastern Africa /Southern Africa / Western Africa /Central Africa)

### Three types of analyses were considered to analyze the contribution of African scholars to COVID-19 literature

- Analysis of scientific production and research collaboration by geographical region As an introductory step to a better understanding of the global COVID-19 research, we quantified absolute scientific production by regions by counting the number of documents authored by researchers from each region. Moreover, we compared inter-regional, and international collaborations. We also compared the research leadership. The concepts used in the present study are defined as follows:
  – International collaboration: joint participation in the authorship of a document by researchers from two or more countries.
  – Inter-regional collaboration: joint participation in the authorship of a document by researchers from countries in two or more regions.
  Geographical location of authors were taken from authors’ institutional affiliations. The limitations section of this paper includes an in-depth explanation of shortcomings which should be considered when interpreting the results.
- Analysis of research production, collaboration and leadership from researchers residing in countries in Africa To specifically analyze COVID-19 research publications from African countries, we determined the number of documents authored by researchers from these countries. Furthermore, a direct collaboration network, representing the main African countries collaborating in COVID-19 research, was generated.
- Subject areas and research fields in global COVID-19 research production We analyzed the research subject areas and fields according to the disciplines that contributed the most to scientific production on COVID-19, as identified by means of the subject area classification of scientific journals in the WoS Core Collection (WoS-CC) as well as the keywords plus descriptors and qualifiers assigned to the documents. To compare research orientations, we presented data for global research output, for publications produced solely by researchers from African countries, and publications produced through collaborations between researchers from African and non-African countries and territories. Data analyses to extract publication indicators were performed using Excel, Biblioshiny package of the R software (16).

## Results

### African scientific production by region and degree of international collaborations

Considering African participation to the scientific production related to COVID-19, Northern Africa (34.07%) and Southern Africa (31.49%) are the main contributors (Table 1). Together, these regions contributed up to 64.55% of the African scientific research production on COVID-19 that was indexed in the consulted sources. Central Africa contributed the least, only 5% of the African scientific production (Table 1). Amongst these scientific collaborations and partnerships, 41.23% of the scholarly research output was conducted by a country without collaboration with other African or non-African countries. This scientific production trend contrasts with the high percentages of collaborations observed in some specific African regions: namely, in Central Africa, 83.58% of the papers were published in collaboration with authors from more than one country, in Southern Africa 63.41%, and in Northern Africa, 57.20%.

**Table 1:** Scientific production on COVID-19, broken down by geographical region

| Geographical area | Total documents |  | Eastern Africa |  | Southern Africa |  | Western Africa |  | Central Africa |  | Northern Africa |  |
| --- | --- | --- | --- | --- | --- | --- | --- | --- | --- | --- | --- | --- |
|  | N | % | N | % | N | % | N | % | N | % | N | % |
| North America | 1,298 | 24.20 | 314 | 31.09 | 529 | 31.32 | 323 | 27.01 | 83 | 30.97 | 353 | 19.32 |
| Latin America & the Caribbean | 406 | 7.57 | 99 | 9.80 | 198 | 11.72 | 113 | 9.45 | 23 | 8.58 | 129 | 7.06 |
| Europe & Central Asia | 1,825 | 34.03 | 391 | 38.71 | 715 | 42.33 | 456 | 38.13 | 162 | 60.45 | 522 | 28.57 |
| East Asia & Pacific | 868 | 16.18 | 188 | 18.61 | 337 | 19.95 | 252 | 21.07 | 47 | 17.54 | 250 | 13.68 |
| Sub-Saharan Africa | 3,660 | 68.25 | 1010 | 100.00 | 1689 | 100.00 | 1196 | 100.00 | 268 | 100.00 | 126 | 6.90 |
| Eastern Africa ** | 1,010 | 18.83 | 1,010 | 100.00 | 158 | 9.35 | 133 | 11.12 | 49 | 18.28 | 55 | 3.01 |
| Southern Africa** | 1,689 | 31.49 | 158 | 15.64 | 1689 | 100.00 | 165 | 13.80 | 59 | 22.01 | 54 | 2.96 |
| Western Africa** | 1196 | 22.30 | 133 | 13.17 | 165 | 9.77 | 1196 | 100.00 | 65 | 24.25 | 63 | 3.45 |
| Central Africa** | 268 | 5.00 | 49 | 4.85 | 59 | 3.49 | 65 | 5.43 | 268 | 100.00 | 12 | 0.66 |
| Middle East & North Africa | 2068 | 38.56 | 124 | 12.28 | 179 | 10.60 | 155 | 12.96 | 24 | 8.96 | 1827 | 100.00 |
| Middle East* | 779 | 14.53 | 99 | 9.80 | 148 | 8.76 | 123 | 10.28 | 14 | 5.22 | 538 | 29.45 |
| Northern Africa* | 1827 | 34.07 | 55 | 5.45 | 54 | 3.20 | 63 | 5.27 | 12 | 4.48 | 1827 | 100.00 |
| South Asia | 471 | 8.78 | 122 | 12.08 | 157 | 9.30 | 134 | 11.20 | 26 | 9.70 | 183 | 10.02 |
| Inter-regional collaboration | 134 | 2.50 | 48 | 4.75 | 83 | 4.91 | 73 | 6.10 | 22 | 8.21 | 17 | 0.93 |
| International collaboration | 3019 | 56.29 | 617 | 61.09 | 988 | 58.50 | 697 | 58.28 | 202 | 75.37 | 1029 | 56.32 |
| no or national collaboration | 2211 | 41.23 | 345 | 34.16 | 618 | 36.59 | 427 | 35.70 | 44 | 16.42 | 782 | 42.80 |
| **Total** | **5,363** | **100** | **1,010** | 100.00 | **1689** | 100.00 | **1196** | 100.00 | **268** | 100.00 | **1827** | 100.00 |

\*\* Sub-region of the Sub-Saharan Africa \* Sub-region of the Middle East & North Africa

Europe, Central Asia and North America based researchers are the main collaborators of African researchers, representing respectively 34.03% and 24.20% of scientific partnership contributing to the production related to COVID-19 (Table 1).

Northern Africa researchers collaborated in less than 4% of their production with other African regions. Their main collaborators are from the Middle East and Europe & Central Asia researchers, with respectively 29.45% and 28.57% of scientific output related to COVID-19 (Table 1).

Within Africa, Central Africa researchers mostly collaborated with Western Africa (24.25%), followed by Southern Africa (22.01%). Outside Africa, we observed that Europe & Central Asia researchers were their principal collaborators (60.45%), followed by Northern America (30.97%) (Table 1).

Concerning Western Africa researchers, they mostly collaborated within Africa with Southern Africa (13.80%), follow by Eastern Africa (11.12%); while Europe & Central Asia researchers (38.13%), followed by Northern America (27.01%) were their principal collaborators outside Africa (Table 1).

Southern Africa researchers mostly collaborated in Africa with Western Africa and Eastern Africa combined in less than 10% of their production. Europe & Central Asia researchers (42.33%), followed by Northern America (31.32%) were their principal collaborators outside Africa (Table 1).

Eastern Africa researchers collaborated with scholars residing in Africa mostly with Southern Africa (15.64%), followed by Western Africa (13.17%). Europe & Central Asia researchers (38.71%), followed by Northern America (31.09%) were their principal collaborators outside Africa (Table 1).

Figure 2, show the strength of COVID-19 research collaboration networks between African and other institutions. The diameter of the circles and color codes represent the Spearman value of correlation coefficients. The larger (or the lower) the value, the higher (or the lower) the collaboration strength between regions. The Figure shows a very weak correlation between researchers from Sub-Saharan Africa and Northern Africa.

**Figure 2:**
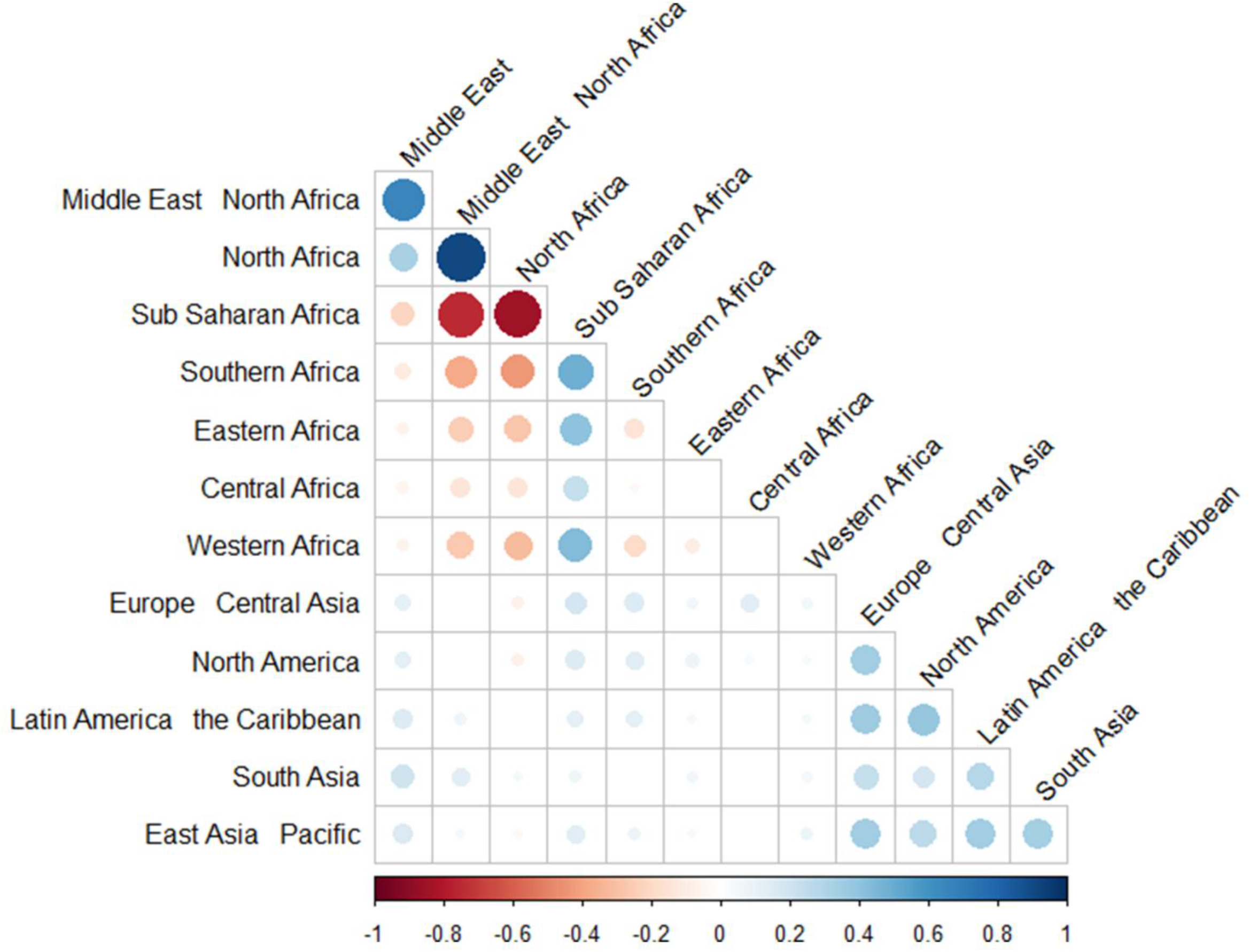
Correlation heatmap denoting the strength of COVID-19 research collaboration networks between African and other institutions. The diameter of the circles and color codes represent the Spearman value of correlation coefficients. The larger (or the lower) the value, the higher (or the lower) the collaboration strength between regions.

### Scientific papers published by country and degree of international collaborations

Research production in Africa is concentrated in South Africa and Egypt, whose researchers contributed respectively to 27.07% and 22.75% of the documents from their regions. These countries are followed by : Nigeria (14.12%), Morocco (6.82%), Ethiopia (6.00%) and Kenya (5.39%).

A total of fifty-two countries contributed to Africa’s scientific production, with the number of documents by country ranging from 2 to 1,452; the mean number of documents per country was 123.75. In Central Africa, the country with the highest contribution was Cameroon with 127 (2.3%) documents, while Ethiopia leads the production in the Eastern East with 322 (6.00%) articles, Egypt in the North Africa with 1220 (22.75%) documents, South Africa in the Southern Africa with 1452 (27.07%) articles and Nigeria in the Western Africa with 757 (14.12%) items (Table 2).

**Table 2:** Africa scientific production on COVID-19, by country

| PAYS | World Bank FY21 analytical classifications by region | Total documents |  | No collaboration |  | International collaborations |  |
| --- | --- | --- | --- | --- | --- | --- | --- |
|  |  | N | % | NO | % | N | % |
| South Africa | Southern Africa | 1452 | 27.07 | 559 | 38.50 | 893 | 61.50 |
| Egypt | North Africa | 1220 | 22.75 | 405 | 33.20 | 815 | 66.80 |
| Nigeria | Western Africa | 757 | 14.12 | 310 | 40.95 | 447 | 59.05 |
| Morocco | North Africa | 366 | 6.82 | 248 | 67.76 | 118 | 32.24 |
| Ethiopia | Eastern Africa | 322 | 6.00 | 196 | 60.87 | 126 | 39.13 |
| Kenya | Eastern Africa | 289 | 5.39 | 53 | 18.34 | 236 | 81.66 |
| Ghana | Western Africa | 234 | 4.36 | 68 | 29.06 | 166 | 70.94 |
| Uganda | Eastern Africa | 169 | 3.15 | 31 | 18.34 | 138 | 81.66 |
| Tunisia | North Africa | 159 | 2.96 | 59 | 37.11 | 100 | 62.89 |
| Cameroon | Central Africa | 127 | 2.37 | 28 | 22.05 | 99 | 77.95 |
| Algeria | North Africa | 113 | 2.11 | 43 | 38.05 | 70 | 61.95 |
| Sudan | Eastern Africa | 113 | 2.11 | 30 | 26.55 | 83 | 73.45 |
| Zimbabwe | Southern Africa | 91 | 1.70 | 28 | 30.77 | 63 | 69.23 |
| Tanzania | Eastern Africa | 89 | 1.66 | 13 | 14.61 | 76 | 85.39 |
| Senegal | Western Africa | 88 | 1.64 | 20 | 22.73 | 68 | 77.27 |
| D. R. Congo | Central Africa | 81 | 1.51 | 11 | 13.58 | 70 | 86.42 |
| Mozambique | Southern Africa | 65 | 1.21 | 1 | 1.54 | 64 | 98.46 |
| Malawi | Southern Africa | 57 | 1.06 | 8 | 14.04 | 49 | 85.96 |
| Zambia | Southern Africa | 57 | 1.06 | 5 | 8.77 | 52 | 91.23 |
| Libya | North Africa | 56 | 1.04 | 27 | 48.21 | 29 | 51.79 |
| Rwanda | Eastern Africa | 51 | 0.95 | 4 | 7.84 | 47 | 92.16 |
| Congo | Central Africa | 43 | 0.80 | 3 | 6.98 | 40 | 93.02 |
| Mali | Western Africa | 41 | 0.76 | 3 | 7.32 | 38 | 92.68 |
| Burkina Faso | Western Africa | 35 | 0.65 | 9 | 25.71 | 26 | 74.29 |
| Mauritius | Eastern Africa | 32 | 0.60 | 10 | 31.25 | 22 | 68.75 |
| Sierra Leone | Western Africa | 31 | 0.58 | 3 | 9.68 | 28 | 90.32 |
| Botswana | Southern Africa | 29 | 0.54 | 9 | 31.03 | 20 | 68.97 |
| Madagascar | Eastern Africa | 29 | 0.54 | 6 | 20.69 | 23 | 79.31 |
| Benin | Western Africa | 27 | 0.50 | 3 | 11.11 | 24 | 88.89 |
| The Gambia | Western Africa | 23 | 0.43 | 4 | 17.39 | 19 | 82.61 |
| Gabon | Central Africa | 22 | 0.41 | 2 | 9.09 | 20 | 90.91 |
| Guinea | Western Africa | 21 | 0.39 | 3 | 14.29 | 18 | 85.71 |
| Ivory Coast | Western Africa | 21 | 0.39 | 0 | 0.00 | 21 | 100.00 |
| Namibia | Southern Africa | 17 | 0.32 | 6 | 35.29 | 11 | 64.71 |
| Niger | Western Africa | 15 | 0.28 | 3 | 20.00 | 12 | 80.00 |
| Somalia | Eastern Africa | 12 | 0.22 | 2 | 16.67 | 10 | 83.33 |
| Swaziland | Southern Africa | 11 | 0.21 | 5 | 45.45 | 6 | 54.55 |
| Togo | Western Africa | 10 | 0.19 | 0 | 0.00 | 10 | 100.00 |
| Liberia | Western Africa | 9 | 0.17 | 2 | 22.22 | 7 | 77.78 |
| Guinea-Bissau | Western Africa | 6 | 0.11 | 0 | 0.00 | 6 | 100.00 |
| Mauritania | Western Africa | 6 | 0.11 | 0 | 0.00 | 6 | 100.00 |
| Burundi | Eastern Africa | 5 | 0.09 | 0 | 0.00 | 5 | 100.00 |
| Central African Republic | Central Africa | 5 | 0.09 | 0 | 0.00 | 5 | 100.00 |
| Chad | Central Africa | 5 | 0.09 | 0 | 0.00 | 5 | 100.00 |
| Eritrea | Eastern Africa | 4 | 0.07 | 0 | 0.00 | 4 | 100.00 |
| South Sudan | Eastern Africa | 4 | 0.07 | 1 | 25.00 | 3 | 75.00 |
| Angola | Southern Africa | 3 | 0.06 | 3 | 100.00 | 0 | 0.00 |
| Djibouti | North Africa | 3 | 0.06 | 1 | 33.33 | 2 | 66.67 |
| Equatorial Guinea | Central Africa | 3 | 0.06 | 0 | 0.00 | 3 | 100.00 |
| Lesotho | Southern Africa | 3 | 0.06 | 1 | 33.33 | 2 | 66.67 |
| Comoros | Eastern Africa | 2 | 0.04 | 0 | 0.00 | 2 | 100.00 |
| Seychelles | Eastern Africa | 2 | 0.04 | 0 | 0.00 | 2 | 100.00 |

Among the most productive countries (> 50 documents), Morocco, Ethiopia, Libya, Nigeria, and South Africa presented the lowest proportion of international collaborations. However, many other countries showed a value of international collaborations that exceed 80% (Table 2).

### African and Non-African countries collaboration and the impact of their research

Table S1 contains data on the collaborations between researchers in Africa and those abroad. African research output on COVID-19 is characterized by its cooperative links, particularly with the USA and UK which collaborated respectively with 49 and 45 African countries. We observed a significant number of links for colonial countries (Table 3, Table S1).

**Table 3:** Collaboration and leadership of top 20 African countries in research papers on COVID-19

| Rank | Country | Colonial country | Total Collaborations |  |  |  |  |  | Collaborations with African countries |  |  |  | Collaborations with non-African countries |  |  |  |
| --- | --- | --- | --- | --- | --- | --- | --- | --- | --- | --- | --- | --- | --- | --- | --- | --- |
|  |  |  | N countries | N collaborations | N collaborations (%) | Average citations per item | h-index | Main countries collaborators (n collaborations) | N countries | Average citations per item | h-index | Main African collaborators (n collaborations) | N countries | Average citations per item | h-index | Main non-African collaborators (n collaborations) |
| 0 | ALL | / | 173 | 5363 | 100 | 4.15 | 57 | South Africa (n = 1156); Egypt (n = 1220); USA (n = 1156) | 48 | 2.2 | 28 | South Africa (n = 636); Egypt (n = 413); Nigeria (n = 350) | 119 | 5.7 | 53 | USA (n = 1156); UK (n = 955); South Africa (n = 816) |
| 1 | South Africa | UK | 138 | 893 | 61.50 | 5.80 | 33.00 | USA (n = 414); UK (n = 360); Australia (n = 158) | 43 | 2.42 | 13 | Nigeria (n = 97); Kenya (n = 62); Ghana (n = 39) | 95 | 8.53 | 33 | USA (n = 414); UK (n = 360); Australia (n = 158) |
| 2 | Egypt | UK | 128 | 815 | 66.80 | 4.89 | 31.00 | Saudi Arabia (n = 317); USA (n = 257); UK (n = 172) | 28 | 3.81 | 15 | Nigeria (n = 36); South Africa (n = 35); Tunisia (n = 28) | 100 | 5.43 | 27 | Saudi Arabia (n = 317); USA (n = 257); UK (n = 172) |
| 3 | Nigeria | UK | 125 | 447 | 59.05 | 3.93 | 19.00 | UK (n = 175); USA (n = 159); South Africa (n = 97) | 38 | 1.91 | 12 | South Africa (n = 97); Egypt (n = 43); Egypt (n = 36) | 87 | 5.59 | 17 | UK (n = 175); USA (n = 159); India (n = 68) |
| 4 | Morocco | France | 92 | 118 | 32.24 | 2.97 | 15.00 | France (n = 33); USA (n = 32); Saudi Arabia (n = 24) | 24 | 2.46 | 9 | Egypt (n = 21); Algeria (n = 18); Tunisia (n = 12) | 68 | 4.15 | 10 | France (n = 33); USA (n = 32); Saudi Arabia (n = 24) |
| 5 | Ethiopia | / | 77 | 126 | 39.13 | 3.94 | 16 | USA (n = 48); UK (n = 32); India (n = 24) | 28 | 2.76 | 10 | Nigeria (n = 19); Kenya (n = 14); South Africa (n = 13) | 49 | 6.29 | 12 | USA (n = 48); UK (n = 32); India (n = 24) |
| 6 | Kenya | UK | 116 | 236 | 81.66 | 3.89 | 15.00 | USA (n = 114); UK (n = 98); Canada (n = 34) | 40 | 2.2 | 6 | South Africa (n = 62); Nigeria (n = 43); Uganda (n = 29) | 76 | 4.38 | 13 | USA (n = 114); UK (n = 98); Canada (n = 34) |
| 7 | Ghana | UK | 97 | 166 | 70.94 | 2.92 | 12.00 | UK (n = 68); USA (n = 60); South Africa (n = 39) | 34 | 1.24 | 4 | South Africa (n = 39); Nigeria (n = 32); Kenya (n = 19) | 63 | 3.91 | 11 | UK (n = 68); USA (n = 60); Germany (n = 29) |
| 8 | Uganda | UK | 101 | 138 | 81.66 | 3.64 | 12.00 | USA (n = 70); UK (n = 55); South Africa (n = 31) | 29 | 3.66 | 4 | South Africa (n = 31); Kenya (n = 29); Nigeria (n = 21) | 72 | 3.63 | 12 | USA (n = 70); UK (n = 55); Canada (n = 20) |
| 9 | Tunisia | France | 89 | 100 | 62.89 | 6.03 | 16 | USA (n = 39); Saudi Arabia (n = 33); Italy (n = 30) | 17 | 2.43 | 4 | Egypt (n = 28); Nigeria (n = 16); Morocco (n = 12) | 72 | 8.2 | 16 | USA (n = 39); Saudi Arabia (n = 33); Italy (n = 30) |
| 10 | Cameroon | UK /France | 87 | 99 | 77.95 | 4.85 | 10.00 | USA (n = 36); France (n = 33); UK (n = 27) | 36 | 2.08 | 3 | South Africa (n = 19); Kenya (n = 13); Ghana (n = 13) | 51 | 6.35 | 9 | USA (n = 36); France (n = 33); UK (n = 27) |
| 11 | Sudan | UK | 78 | 83 | 73.45 | 5.42 | 9.00 | UK (n = 32); Saudi Arabia (n = 32); Egypt (n = 19) | 23 | 3 | 4 | Egypt (n = 19); Nigeria (n = 11); South Africa (n = 11) | 55 | 6.14 | 9 | UK (n = 32); Saudi Arabia (n = 32); USA (n = 15) |
| 12 | Algeria | France | 81 | 70 | 61.95 | 2.16 | 8.00 | Saudi Arabia (n = 19); Egypt (n = 18); Morocco<br>(n = 18) | 15 | 1.78 | 4 | Egypt (n = 18); Morocco (n = 18); Tunisia (n = 11) | 66 | 2.42 | 7 | Saudi Arabia (n = 19); France (n = 16); USA<br>(n = 15) |
| 13 | Zimbabwe | UK | 89 | 63 | 69.23 | 3.77 | 9.00 | South Africa<br>(n = 32); UK<br>(n = 29); USA<br>(n = 23) | 28 | 1.86 | 4 | South Africa<br>(n = 32); Kenya<br>(n = 11); Uganda<br>(n = 10) | 61 | 5.36 | 8 | UK (n = 29); USA<br>(n = 23); Canada<br>(n = 8) |
| 14 | Tanzania | UK | 80 | 76 | 85.39 | 4.91 | 11.00 | UK (n = 32); USA<br>(n = 29); South<br>Africa (n = 17) | 25 | 4.17 | 4 | South Africa<br>(n = 17); Uganda<br>(n = 17); Nigeria<br>(n = 14) | 55 | 5.14 | 9 | UK (n = 32); USA<br>(n = 29);<br>Australia (n = 10) |
| 15 | Senegal | France | 68 | 68 | 77.27 | 14.24 | 13.00 | USA (n = 28);<br>France (n = 22);<br>UK (n = 22) | 30 | 0.46 | 2 | South Africa<br>(n = 14); Nigeria<br>(n = 9);<br>Cameroon (n = 8) | 38 | 20.6 | 13 | USA (n = 28);<br>France (n = 22);<br>UK (n = 22) |
| 16 | Democratic<br>Republic of<br>the Congo | Belgium | 56 | 70 | 86.42 | 3.6 | 7.00 | Belgium (n = 30);<br>USA (n = 26); UK<br>(n = 23) | 27 | 2.08 | 4 | South Africa<br>(n = 19); Kenya<br>(n = 9);<br>Cameroon (n = 7) | 29 | 3.96 | 7 | Belgium (n = 30);<br>USA (n = 26); UK<br>(n = 23) |
| 17 | Mozambique | Portugal | 85 | 64 | 98.46 | 16.29 | 10.00 | UK (n = 29);<br>Spain (n = 28);<br>USA (n = 20) | 24 | 0 | 0 | South Africa<br>(n = 11); Uganda<br>(n = 8); Tanzania<br>(n = 5) | 61 | 16.64 | 10 | UK (n = 29);<br>Spain (n = 28);<br>USA (n = 20) |
| 18 | Zambia | UK | 77 | 52 | 91.23 | 4.32 | 6.00 | USA (n = 30); UK<br>(n = 22); South<br>Africa (n = 13) | 27 | 0.8 | 1 | South Africa<br>(n = 13); Kenya<br>(n = 10); Uganda<br>(n = 9) | 50 | 4.74 | 6 | USA (n = 30); UK<br>(n = 22); China<br>(n = 10) |
| 19 | Malawi | UK | 61 | 49 | 85.96 | 4.60 | 10.00 | UK (n = 31);<br>South Africa<br>(n = 15); USA<br>(n = 14) | 25 | 1 | 1 | South Africa<br>(n = 15); Kenya<br>(n = 11); Nigeria<br>(n = 8) | 36 | 5.57 | 9 | UK (n = 31); USA<br>(n = 14); Sweden<br>(n = 8) |
| 20 | Libya | Italy | 67 | 29 | 51.79 | 3.06 | 6.00 | UK (n = 18);<br>Saudi Arabia<br>(n = 10); Egypt<br>(n = 9) | 7 | 2.83 | 5 | Egypt (n = 9);<br>Nigeria (n = 5);<br>Kenya (n = 4) | 60 | 3.29 | 5 | UK (n = 18);<br>Saudi Arabia<br>(n = 10); Italy<br>(n = 8) |

Concerning collaborations between African countries, South Africa stands out for its strong intra-regional ties, and it has become the main reference for research collaboration on COVID-19, both in Africa and among the top 20 most productive African countries. It has collaborated with 43 different African countries (Table 3, Table S2). Kenya ranks second in terms of collaborative leadership within Africa, followed by Nigeria and Cameroon which collaborated respectively with 40, 38 and 36 other African countries (Table 3, Table S2). On the other hand, Egypt was the second main contributor of scientific production but it only collaborated with researchers of 28 African countries; it was however the main collaborators of Northern Africa countries. Egypt’s principal collaborators was Saudi Arabia, followed by the USA. It is also important to mention that Saudi Arabia was among the main collaborator of other North Africa countries (in particular, Arab speaking countries) (Table 3, Table S1).

The published articles considered in our analysis had an average citation per item of 4.15 and h-index of 57. These scores were higher in the scientific production in collaboration with non-African researchers, when compared to solely African collaboration, with respective 5.7 vs 2.2 for average citations per item and 53 vs 28 for h-index (Table 3).

### Active journals

*Pan African Medical Journal, South African Medical Journal, PLoS ONE, BMJ Global Health* and *Journal of Biomolecular Structure and Dynamics* was the top 5 leading journals with respectively 246 (4.59%), 155 (2.89%), 97 (1.813%), 59 (1.10%) and 47 (0.88%) documents. In the list of top 15 active journals worldwide, two journals were in the field of microbiology and infections while the remaining were in the field of public health, environment, and general medicine (Table 4). The mean of impact factor of these top 15 journals was 6.26 with and standard deviation of 14.49 and median of 2.74.

**Table 4:** Top 15 active journals publishing research papers on COVID-19 in Africa

|  | Global publications |  |  |  | Solely African publications |  |  |  | African + global collaborations |  |  |  |
| --- | --- | --- | --- | --- | --- | --- | --- | --- | --- | --- | --- | --- |
|  | Journal | N | % | IF | Journal | N | % | IF | Journal | N | % | IF |
| 1 | Pan African Medical Journal | 246 | 4.59 | 0.51 | Pan African Medical Journal | 181 | 7.72 | 0.51 | Pan African Medical Journal | 65 | 2.15 | 0.51 |
| 2 | South African Medical Journal | 155 | 2.89 | 1.70 | South African Medical Journal | 138 | 5.89 | 1.70 | BMJ Global Health | 53 | 1.76 | 4.28 |
| 3 | PLoS ONE | 97 | 1.81 | 2.74 | PLoS ONE | 51 | 2.18 | 2.74 | PLoS ONE | 46 | 1.52 | 2.74 |
| 4 | BMJ Global Health | 59 | 1.10 | 4.28 | African Journal of Primary Health Care and Family Medicine | 27 | 1.15 | 0.93 | Lancet | 40 | 1.32 | 60.39 |
| 5 | Journal of Biomolecular Structure and Dynamics | 47 | 0.88 | 3.55 | Risk Management and Healthcare Policy | 24 | 1.02 | 2.84 | International Journal of Environmental Research and Public Health | 38 | 1.26 | 2.85 |
| 6 | Lancet | 46 | 0.86 | 60.39 | Egyptian Journal of Radiology and Nuclear Medicine | 23 | 0.98 | 0.29 | International Journal of Infectious Diseases | 32 | 1.06 | 3.20 |
| 7 | International Journal of Infectious Diseases | 45 | 0.84 | 3.20 | Journal of Biomolecular Structure and Dynamics | 22 | 0.94 | 3.55 | Journal of Global Health | 26 | 0.86 | 2.90 |
| 8 | Journal of Medical Virology | 45 | 0.84 | 2.02 | Medical Hypotheses | 21 | 0.90 | 1.38 | American Journal of Tropical Medicine and Hygiene | 51 | 1.69 | 2.13 |
| 9 | International Journal of Environmental Research and Public Health | 40 | 0.75 | 2.85 | Journal of Medical Virology | 21 | 0.90 | 2.02 | Journal of Biomolecular Structure and Dynamics | 25 | 0.83 | 3.55 |
| 10 | Medical Hypotheses | 34 | 0.63 | 1.38 | Infection and Drug Resistance | 18 | 0.77 | 2.98 | BMJ-British Medical Journal | 17 | 0.56 | 30.31 |
| 11 | American Journal of Tropical Medicine and Hygiene | 64 | 1.19 | 2.13 | HTS Teologiese Studies / Theological Studies | 17 | 0.73 | 0.52 | Journal of Medical Virology | 24 | 0.79 | 2.02 |
| 12 | South African Medical Journal | 33 | 0.62 | 1.29 | South African Journal of Science | 17 | 0.73 | 1.70 | Frontiers in Public Health | 24 | 0.79 | 2.13 |
| 13 | Journal of Global Health | 32 | 0.60 | 2.90 | Heliyon | 16 | 0.68 | 1.86 | Travel Medicine and Infectious Disease | 22 | 0.73 | 4.59 |
| 14 | Frontiers in Public Health | 31 | 0.58 | 2.13 | International Journal of Infectious Diseases | 13 | 0.55 | 3.20 | The Lancet Global Health | 21 | 0.70 | 21.60 |
| 15 | Risk Management and Healthcare Policy | 30 | 0.56 | 2.84 | Pharmacy Education | 13 | 0.55 | 0.30 | Clinical Infectious Diseases | 18 | 0.60 | 8.31 |

Comparing the contribution of solely African researchers and those in collaboration with non-African researchers, the average impact factor of the top 15 journals was about six times higher in the group of researchers who collaborated with non-African researchers at 10.09 versus 1.77, with medians of 3.20 versus 1.70.

### Subject areas addressed in publications on COVID-19 in Africa

The analysis on scientific COVID-19 output, produced by all countries worldwide, by African countries alone, and through Africa plus global collaborations, showed differences in terms of disciplinary orientations and research topics. In terms of disciplines involved, discordance was noted between global publications versus solely African publications (Table 5). There is also certain discordance between solely African publications and Africa plus global collaborations. In contrast, there is great affinity between global research output and output from Africa plus global collaborations. Of note, COVID-19 publications from Africa alone and from Africa plus global collaborations were dominated by papers in the field of “Public, Environmental & Occupational Health,” and “Infectious Diseases” although the proportions are slightly higher from Africa plus global collaborations. The disciplines of “Medicine, General & Internal” and “Health Policy & Services” were also of great relevance in the publications from African countries alone (Table 5).

**Table 5:** COVID-19 related research papers broken down by Web of Science categories, according to African involvement

| Rank | WoS Category | Global publications |  | Solely African publications |  | African + global collaborations |  |
| --- | --- | --- | --- | --- | --- | --- | --- |
|  |  | N | % | N | % | N | % |
| 1 | Public. Environmental & Occupational Health | 1013 | 22.70 | 438 | 22.20 | 575 | 23.11 |
| 2 | Infectious Diseases | 547 | 12.26 | 207 | 10.49 | 340 | 13.67 |
| 3 | Medicine. General & Internal | 404 | 9.05 | 201 | 10.19 | 203 | 8.16 |
| 4 | Health Care Sciences & Services | 249 | 5.58 | 123 | 6.23 | 126 | 5.06 |
| 5 | Pharmacology & Pharmacy | 225 | 5.04 | 91 | 4.61 | 134 | 5.39 |
| 6 | Biochemistry & Molecular Biology | 174 | 3.90 | 62 | 3.14 | 112 | 4.50 |
| 7 | Multidisciplinary Sciences | 173 | 3.88 | 82 | 4.16 | 91 | 3.66 |
| 8 | Immunology | 163 | 3.65 | 58 | 2.94 | 105 | 4.22 |
| 9 | Respiratory System | 156 | 3.50 | 65 | 3.29 | 91 | 3.66 |
| 10 | Environmental Sciences | 148 | 3.32 | 41 | 2.08 | 107 | 4.30 |
| 11 | Medicine, Research & Experimental | 143 | 3.20 | 71 | 3.60 | 72 | 2.89 |
| 12 | Microbiology | 130 | 2.91 | 51 | 2.58 | 79 | 3.18 |
| 13 | Virology | 128 | 2.87 | 53 | 2.69 | 75 | 3.01 |
| 14 | Pediatrics | 99 | 2.22 | 42 | 2.13 | 57 | 2.29 |
| 15 | Health Policy & Services | 99 | 2.22 | 60 | 3.04 | 39 | 1.57 |
| 16 | Clinical Neurology | 95 | 2.13 | 34 | 1.72 | 61 | 2.45 |
| 17 | Surgery | 95 | 2.13 | 46 | 2.33 | 49 | 1.97 |
| 18 | Tropical Medicine | 81 | 1.82 | 29 | 1.47 | 52 | 2.09 |
| 19 | Oncology | 81 | 1.82 | 37 | 1.88 | 44 | 1.77 |
| 20 | Psychiatry | 79 | 1.77 | 35 | 1.77 | 44 | 1.77 |

## Discussion

In the present bibliometric study, we found that COVID-19 related collaboration patterns varied among African regions, being shaped and driven by historical, social, cultural, linguistic, and even religious determinants. For instance, most of the scholarly partnerships occurred with formerly colonial countries (like European or North-American countries). In other cases, scholarly ties of North African countries were above all with the Kingdom of Saudi Arabia. In terms of amount of publications, South Africa and Egypt were among the most productive countries.

Bibliometrics and, in particular, scientometrics can help scholars identify research areas of particular interest, as well as emerging topics. Moreover, they can assist decision- and policy-makers in allocating funding and economic-financial, logistic, organizational, and human resources, based on the specific gaps and needs of a given country or research area.

Several, important initiatives such as the “Hinari Access to Research for Health Programme” (HINARI) established by the World Health Organization (WHO), involving the scientific community and major publishers, have granted developing countries, including Africa, access to biomedical and health-related scientific literature (17). Recently, the “National Institutes of Health” (NIH) has set up an initiative, termed as UNITE, in order to “*end structural racism and achieve racial equity in the biomedical research enterprise*”. Despite these efforts, the contribution of African countries to global knowledge has decreased in the last years in terms of their share.

A study (18) explored public health-related investigations conducted by African scholars in the period 1991-2005. An increase in the amount of investigations and international collaborations was reported by 382% and 45-67%, respectively. However, uneven statistics concerning publishing and collaborating trends could be detected, with major regional variations.

Our findings are in line with the existing literature, showing regional differences at the African level. COVID-19 has further distorted and exacerbated some inequalities in publishing and collaborating: for instance, a bibliometric study assessing COVID-19 related papers published in the top 50 medical imaging journals in the period March-May 2020 has found gender-disparities, with women from developing countries being over-represented in low-ranking journals (19). Another study (20), mining the entire COVID-19 related literature, has found that there were fewer women first authors than expected.

In the present study, we found that COVID-19 related publications were mainly focused on topics like “Public, Environmental & Occupational Health”, “Infectious Diseases”, “Medicine, General & Internal” and “Health Policy & Services”. This particular focus can be understood considering that the global burden of disease in African countries is mostly generated by communicable disorders, which can be prevented by implementing public health interventions. It is interesting that in these research topics and fields, African countries as well as other developing countries and territories have performed better with respect to developed countries (21).

As such, we can conclude that the effect of the COVID-19 pandemic is nuanced and complex, on the one hand amplifying already existing inequalities, on the other hand paving the way for new opportunities and catalyzing new venues.

However, despite its strengths, including the methodological rigor, the transparency and reproducibility of the present study, as well as the extensive series of analyses conducted, and the number of electronic scholarly databases mined, this investigation suffers from a number of shortcomings that should be properly acknowledged. Gray literature (via Google Scholar) was not included, as well as other major databases such as Scopus.

## Conclusions

In conclusion, the ongoing COVID-19 pandemic has exerting a subtle, complex impact on research and publishing patterns in African countries. On the one hand, it has distorted and even amplified existing inequalities and disparities in terms of amount of scholarly output, share of global knowledge, and patterns of collaborations, due to the chronic lack of infrastructures, facilities and resources that plagues Africa. In addition, we found that inherited historical, societal, cultural, linguistic, and even religious habits shape the scientific research collaboration in Africa. On the other hand, COVID-19 provided new opportunities for research collaborations, which contributed to generating novel international partners for academic exchanges, and research collaborations. Furthermore, COVID-19 enabled to identify research fields in which African scholars can strengthen their scientific leadership.

## Supporting information

Supplementary for How the COVID-19 pandemic is shaping research in Africa

## Data Availability

All the data is available in the manuscript

## Availability of data and materials

This study is based on publicly available data.

## References

1. Bragazzi NL. Nanomedicine: Insights from a Bibliometrics-Based Analysis of Emerging Publishing and Research Trends.. Medicina (Kaunas). 2019;55.

2. Ghanbari MK, Behzadifar M, Doshmangir L, Martini M, Bakhtiari A, Alikhani M, et al. Mapping Research Trends of Universal Health Coverage From 1990 to 2019: Bibliometric Analysis.. JMIR Public Health Surveill. 2021;7:e24569.

3. Watad A, Bragazzi NL, Adawi M, Amital H, Kivity S, Mahroum N, et al. Is autoimmunology a discipline of its own? A big data-based bibliometric and scientometric analyses. Autoimmunity [Internet]. March 2017;50(4):269–74. Available at: https://doi.org/10.1080%2F08916934.2017.1305361

4. Africa doubles research output over past decade, moves towards a knowledge-based economy Research Trends [Internet]. https://www.researchtrends.com/issue-35-december-2013/africa-doubles-research-output/; Available at: https://www.researchtrends.com/issue-35-december-2013/africa-doubles-research-output/

5. Tijssen RJW. Africa’s contribution to the worldwide research literature: New analytical perspectives trends, and performance indicators. Scientometrics [Internet]. May 2007;71(2):303–27. Available at: https://doi.org/10.1007%2Fs11192-007-1658-3

6. Africa generates less than one percent of the world’s research; data analytics can change that [Internet]. 2018. Available at: https://www.elsevier.com/connect/africa-generates-less-than-1-of-the-worlds-research-data-analytics-can-change-that

7. Confraria H, Godinho MM. The impact of African science: a bibliometric analysis. Scientometrics [Internet]. October 2014;102(2):1241–68. Available at: https://doi.org/10.1007%2Fs11192-014-1463-8

8. Jia Z, Wu Y, Ding F, Wen T. Scientific efforts on SARS-CoV-2 research: A global survey analysis.. J Infect Dev Ctries. 2021;15:185–90.

9. Chen Y, Cheng L, Lian R, Song Z, Tian J. COVID-19 vaccine research focusses on safety, efficacy, immunoinformatics, and vaccine production and delivery: a bibliometric analysis based on VOSviewer.. Biosci Trends. 2021;

10. Farooq RK, Rehman SU, Ashiq M, Siddique N, Ahmad S. Bibliometric analysis of coronavirus disease (COVID-19) literature published in Web of Science 2019-2020.. J Family Community Med. 2021;28:1–7.

11. Ahmad T, Murad MA, Baig M, Hui J. Research trends in COVID-19 vaccine: a bibliometric analysis.. Hum Vaccin Immunother. 2021;:1–6.

12. N VR, Patil SB. Indian Publications on SARS-CoV-2: A bibliometric study of WHO COVID-19 database.. Diabetes Metab Syndr. 2020;14:1171–8.

13. Sachini E, Sioumalas-Christodoulou K, Chrysomallidis C, Siganos G, Bouras N, Karampekios N. COVID-19 enabled co-authoring networks: a country-case analysis.. Scientometrics. 2021;:1–20.

14. Zyoud SH. The Arab region’s contribution to global COVID-19 research: Bibliometric and visualization analysis.. Global Health. 2021;17:31.

15. World Bank Country and Lending Groups – World Bank Data Help Desk [Internet]. https://datahelpdesk.worldbank.org/knowledgebase/articles/906519; Available at: https://datahelpdesk.worldbank.org/knowledgebase/articles/906519

16. Aria M, Cuccurullo C. bibliometrix : An R-tool for comprehensive science mapping analysis. Journal of Informetrics [Internet]. November 2017;11(4):959–75. Available at: https://doi.org/10.1016%2Fj.joi.2017.08.007

17. Hinari Access to Research for Health programme <br> [Internet]. https://www.who.int/hinari/en/; Available at: http://www.who.int/hinari/en/

18. Chuang K-Y, Chuang Y-C, Ho M, Ho Y-S. Bibliometric analysis of public health research in Africa: The overall trend and regional comparisons. South African Journal of Science [Internet]. May 2011;107(5/6). Available at: https://doi.org/10.4102%2Fsajs.v107i5%2F6.309

19. Quak E, Girault G, Thenint MA, Weyts K, Lequesne J, Lasnon C. Author Gender Inequality in Medical Imaging Journals and the COVID-19 Pandemic.. Radiology. 2021;:204417.

20. Andersen JP, Nielsen MW, Simone NL, Lewiss RE, Jagsi R. COVID-19 medical papers have fewer women first authors than expected.. Elife. 2020;9.

21. Wang Q, Zhang C. Can COVID-19 and environmental research in developing countries support these countries to meet the environmental challenges induced by the pandemic?. Environ Sci Pollut Res Int. 2021;

22. Furstenau LB, Rabaioli B, Sott MK, Cossul D, Bender MS, Farina EMJM, et al. A Bibliometric Network Analysis of Coronavirus during the First Eight Months of COVID-19 in 2020.. Int J Environ Res Public Health. 2021;18.

23. Martini M, Bragazzi NL. Googling for Neurological Disorders: From Seeking Health-Related Information to Patient Empowerment, Advocacy, and Open, Public Self-Disclosure in the Neurology 2.0 Era.. J Med Internet Res. 2021;23:e13999.

24. Helliwell JA, Bolton WS, Burke JR, Tiernan JP, Jayne DG, Chapman SJ. Global academic response to COVID-19: Cross-sectional study.. Learn Publ. 2020;

25. International collaboration clusters in Africa [Internet]. https://arxiv.org/ftp/arxiv/papers/1301/1301.5159.pdf; Available at: https://arxiv.org/ftp/arxiv/papers/1301/1301.5159.pdf

