## Supplementary for How the COVID-19 pandemic is shaping research in Africa for "How the COVID-19 pandemic is shaping research in Africa: inequalities in scholarly output and collaborations and new opportunities for scientific leadership"

**Supplementary data 1**

**search terms:**

**African countries and main cities:**

("Algeria" OR "Algerien" OR "Algerie" OR "Algiers" OR "Alger" OR "Angola" OR "Bangui" OR "Republique centrafricaine" OR "Central African Republic" OR "Benin" OR "Dahomey" OR "Benin" OR "Porto-Novo" OR "Botswana" OR "Kalahari" OR "Gaborone" OR "Burkina Faso" OR "Burkina" OR "Ouagadougou" OR "Banfora" OR "Bobo Dioulasso" OR "Burundi" OR "Bujumbura" OR "Gitega" OR "Cameroon" OR "Cameroun" OR "Yaounde" OR "Douala" OR "Bafoussam" OR "Bamenda" OR "Garoua" OR "Maroua" OR "Cape Verde" OR "Cabo Verde" OR "Praia" OR "Chad" OR "Tchad" OR "N'Djamena" OR "Tschad" OR "Comoros" OR "Comores" OR "Congo" OR "Republic of the Congo" OR "Kongo" OR "Brazzaville" OR "Cote d'Ivoire" OR "Ivory Coast" OR "Cote d'Ivoire" OR "Yamoussoukro" OR "Abidjan" OR "Bouake" OR "Korhogo" OR "Democratic Republic of the Congo" OR "Kinshasa" OR "Zaire" OR "Republique democratique du Congo" OR "Djibouti" OR "Egypt" OR "Malawi" OR "Cairo" OR "Al Minya" OR "Equatorial Guinea" OR "Guinee equatoriale" OR "Malabo" OR "Eritrea" OR "Asmara" OR "Ethiopia" OR "Ethiopie" OR "Addis Ababa" OR "Gabon" OR "Gabonese Republic" OR "Libreville" OR "Gambia" OR "Gambie" OR "Banjul" OR "Ghana" OR "Accra" OR "Takoradi" OR "Sekondi" OR "Koforidua" OR "Obuasi" OR "Kumasi" OR "Tamale" OR "Republic of Guinea" OR "Guinea" OR "Guinee" OR "Conakry" OR "Guinea-Bissau" OR "Guine-Bissau" OR "Bissau" OR "Kenya" OR "Kenia" OR "Nairobi" OR "Mombasa" OR "Lesotho" OR "Maseru" OR "Liberia" OR "Republic of Liberia" OR "Liberia" OR "Libya" OR "Libya" OR "Libye" OR "Libia" OR "Madagascar" OR "Madagaskar" OR "Madagasikara" OR "Antananarivo" OR "Toliara" OR "Fianarantsoa" OR "Antsirabe" OR "Toamasina" OR "Mahajanga" OR "Malawi" OR "Malawi" OR "Lilongwe" OR "Mali" OR "Bamako" OR "Sikasso" OR "Koutiala" OR "Koulikoro" OR "Segou" OR "Kayes" OR "Mopti" OR "Mauritania" OR "Mauritanie" OR "Nouakchott" OR "Mauritius" OR "Ile Maurice" OR "Morocco" OR "Morocco" OR "Maroc" OR "Marruecos" OR "Rabat" OR "casablanca" OR "Mozambique" OR "Mosambik" OR "Mocambique" OR "Maputo" OR "Namibia" OR "Namibie" OR "Windhoek" OR "Niger" OR "Niamey" OR "Maradi" OR "Zinder" OR "Tillaberi" OR "Tahoua" OR "Agadez" OR "Nigeria" OR "Abuja" OR "Rwanda" OR "Ruanda" OR "Kigali" OR "Sao Tome" OR "Sao Tome" OR "Senegal" OR "Senegal" OR "Dakar" OR "Ziguinchor" OR "Seychelles" OR "Sierra Leone" OR "Somalia" OR "Somalie" OR "Mogadishu" OR "South Africa" OR "Afrique du Sud" OR "Pretoria" OR "Stellenbosch" OR "Cape Town" OR "Johannesburg" OR "South Sudan" OR "Sudsudan" OR "Juba" OR "Sudan" OR "Soudan" OR "Khartoum" OR "Swaziland" OR "eSwatini" OR "Mbabane" OR "Tanzania" OR "Zanzibar" OR "Tanganyika" OR "Tansania" OR "Tanzanie" OR "Dodoma" OR "Togo" OR "Lome" OR "Tunisia" OR "Tunesien" OR "Tunisie" OR "Tunis" OR "Gabes" OR "Uganda" OR "Ouganda" OR "Kampala" OR "Zambia" OR "Rhodesia" OR "Sambia" OR "Zambie" OR "Lusaka" OR "Zimbabwe" OR "Simbabwe" OR "Harare")

**Terms related to COVID:**

"sars cov 2" OR "covid" OR "COVID 19" OR "COVID 19" OR "ncov" OR "2019 ncov" OR "2019nCoV" OR "novel coronavirus" OR "COVID 19" OR "covid19" OR “covid” OR "new coronavirus" OR "new corona virus" OR "novel corona virus" OR ("Wuhan" AND ("coronavirus" OR "coronavirus" OR "coronaviruses")) OR "sars cov" OR "Coronavirus Disease" OR "SARS-Cov2" OR "Coronavirus 2" OR "2019 coronavirus" OR "coronavirus 2019" OR "Corona Virus Disease" OR "SARS coronavirus" OR "Coronavirus pandemic" OR "coronavirus outbreak*" OR "china corona*" OR "coronavirus" OR "coronaviruses" OR "corona virus"

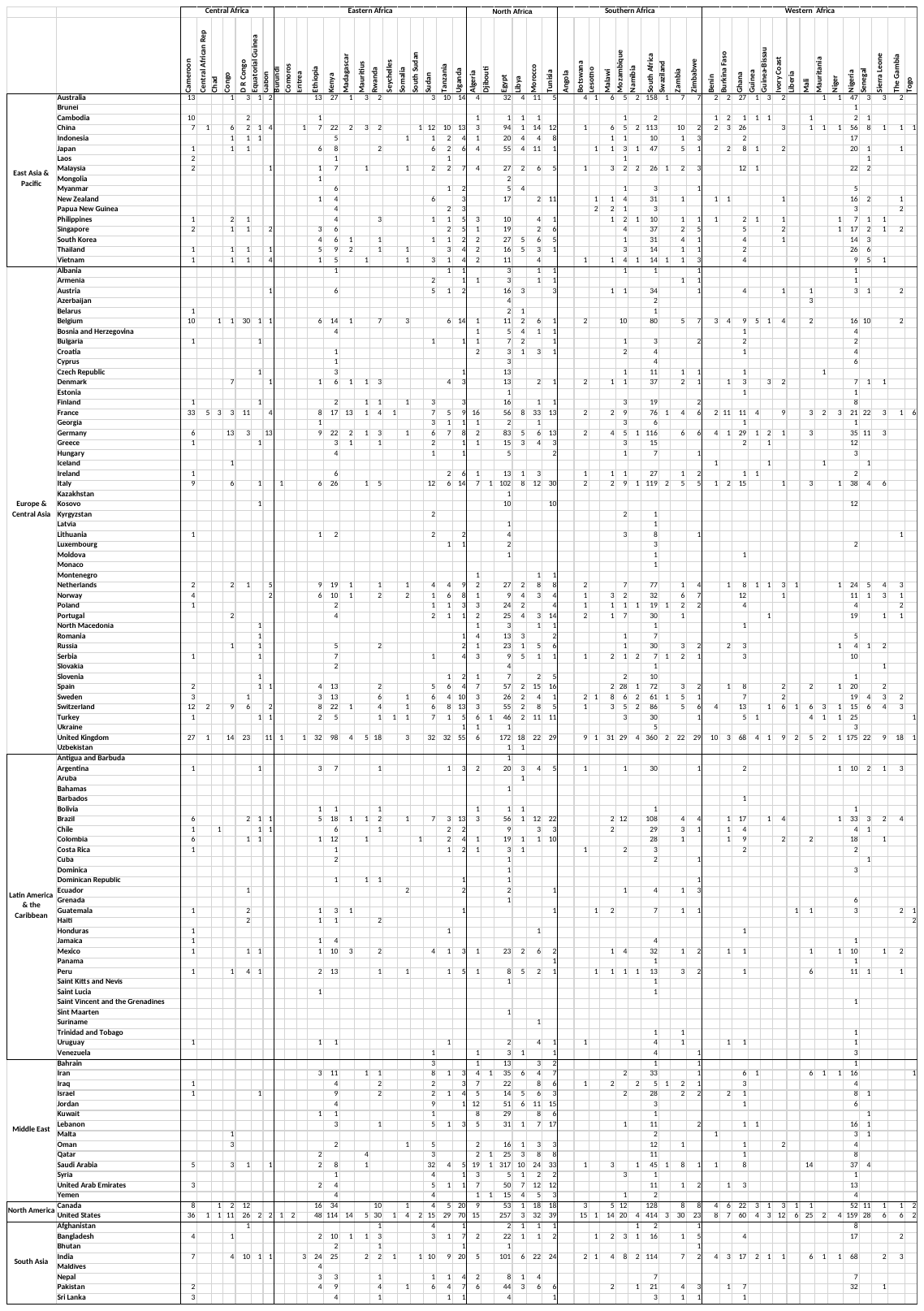

**Table S1:**

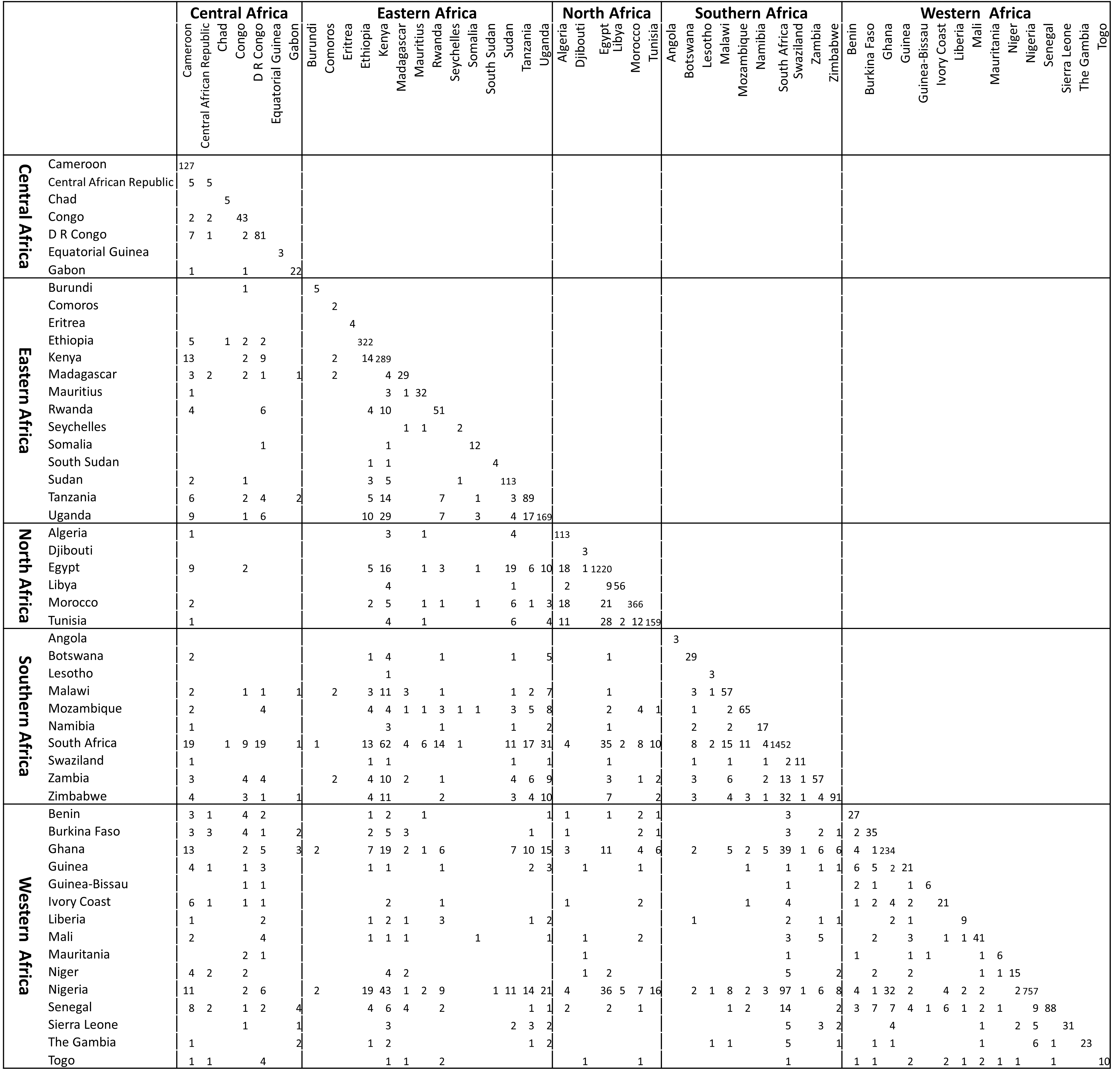

**Table S2:**
